# Simulating the impact of non-pharmaceutical interventions limiting transmission in COVID-19 epidemics using a membrane computing model

**DOI:** 10.1101/2021.07.01.21259828

**Authors:** M Campos, JM Sempere, JC Galán, A Moya, C Llorens, C de-los-Angeles, F Baquero-Artigao, R Cantón, F Baquero

## Abstract

Epidemics caused by microbial organisms are part of the natural phenomena of increasing biological complexity. The heterogeneity and constant variability of hosts, in terms of age, immunological status, family structure, lifestyle, work activities, social and leisure habits, daily division of time, and other demographic characteristics make it extremely difficult to predict the evolution of epidemics. Such prediction is, however, critical for implementing intervention measures in due time and with appropriate intensity. General conclusions should be precluded, given that local parameters dominate the flow of local epidemics. Membrane computing models allows us to reproduce the objects (viruses, hosts) and their interactions (stochastic but also with defined probabilities) with an unprecedented level of detail. Our LOIMOS model helps reproduce the demographics and social aspects of a hypothetical town of 10,320 inhabitants in an average European country where COVID-19 is imported from the outside. The above-mentioned characteristics of hosts and their lifestyle are minutely considered. The dynamics of the epidemics are reproduced and include the effects on viral transmission of innate and acquired immunity at various ages. The model predicts the consequences of delaying the adoption of non-pharmaceutical interventions (between 15 and 45 days after the first reported cases) and the effect of those interventions on infection and mortality rates (reducing transmission by 20%, 50%, and 80%) in immunological response groups. The lockdown for the elderly population as a single intervention appears to be effective. This modelling exercise exemplifies the application of membrane computing for designing appropriate interventions in epidemic situations.

## Introduction

Epidemics are based on an ensemble of transmission events among heterogeneous hosts; in fact, epidemics are determined by the density and intensity of natural biological fluxes among biological entities connected by complex networks that respond to a highly variable ensemble of causes and forces (*Baquero, 2017, Baquero, 2018a*). Membrane-computing differs from conventional mathematical models and most computational models in representing the actors of complex biological scenarios and their multi-hierarchical dynamics and nested interactions as particular computational entities (objects and membranes) (*Păun et al., 2010; Pérez-Jiménez et al., 2003*). In practical terms, cellular membrane computing mimics reality, facilitating the evaluation of possible interventions and decisions to be adopted when facing multilayer challenges, as is the case in complex epidemics.

In the computational model presented in this article, an epidemic process caused by a particular entity (the virus) occurs among computational entities simulating a community with a particular populational structure, contacts, and behaviours resembling those that occur in the real world. This “virtual community approach” has been recently applied to simulate the infective spread of extrachromosomal genetic elements (plasmids) among bacteria and to predict various interventions to limit the spread of antibiotic resistance genes, bacterial species and their clones in the hospital and community settings (*Campos et al 2015; Baquero et al., 2018b; Baquero et al. 2019; Campos et al. 2020; Gil-Gil et al 2021)*

The suitability of this approach for studying the effects of potential interventions aimed at limiting the spread of or perpetuation of COVID-19 epidemics is clear. For this purpose, severe acute respiratory syndrome coronavirus 2 (SARS-CoV-2) is considered a particular object that interacts with other objects (hosts, represented by membranes) in particular scenarios, according to a set of rules and having sensitivity to the frequencies of interacting objects.

Changes in the scenarios, rules and frequencies, resulting from varying interventions, will influence the density of viral-host interactions. These changes are easy to modify in the model, thereby helping to predict the success of each particular intervention or different simultaneous intervention. This task is extremely difficult to test in practice (*Soltesz et al., 2020*) and requires advanced modelling tools. The model applied in this study has significant plasticity; the data entered into this model can be derived from observations of particular settings, such as hospitals, the community, long-term care centres, and schools.

Our simulator (which is also appliable to other viral diseases) is named LOIMOS, from the Ancient Greek λοιμός, meaning plague or pestilence but can refer to any deadly infectious disorder. A user-friendly interface is being developed for LOIMOS and will be freely available. Interested readers should contact our first author, Marcelino Campos.

## MATERIAL AND METHODS

### The basic scenario

First, we start by modelling the dynamics of the epidemic in an isolated COVID-19-free population composed of 10,320 healthy individuals (a number resulting from demographic adjustments) not previously exposed to the causal agent of the infection. After introducing an infected person from another community, three cases were detected in this community. To reflect the natural dynamics of epidemics, our parameters refer to a “wild” model population where no protective measures are considered.

Next, we explored the effect of adopting transmission-decreasing interventions of varying intensity and adopting different times from the onset of the epidemic to evaluate their influence on the epidemiological dynamics. For simplicity’s sake, this article does not detail the particular interventions for reducing transmission; however, the model allows each one of them to be considered, either alone or in combination. These interventions might include 1) the use of surgical masks by the general population, 2) use of FFP2 masks by the general population, 3) the use of facial masks, 4) the use of autonomous respirators in highly exposed health workers, 4) hand washing and disinfection, 5) reduction of sputum formation in the elderly, 6) isolation of COVID-negative elderly individuals at home and nursing care facilities (considered in the study), 7) complete lockdown, i.e., significant limitation in leaving the house, travelling abroad or allowing foreigners to enter the region, 8) partial lockdown, limiting group activities and requiring physical distancing in shops, restaurants, and leisure activities, telework for a portion of the population, and 9) early detection of individuals and contact surveillance for the presence of COVID-19. Given that each of these interventions’ accounts for a percentage of the total reduction in transmission and that their application varies significantly among sites, we decided for this first article to use a global percentage representing the sum of such interventions when applied at different intensities.

#### Parametric structure of the model

The numerical data should correspond to the local data (at any hierarchical level, such as hospitals, towns, regions, nations). Occasionally, the exact quantitative data are not available; therefore, the types of parameters applied can be categorized as 1) assumed parameters, representing choices made a priori based on existing knowledge and scientific literature; 2) measured parameters that will be directly determined from the data, and 3) inferred parameters, which are the quantities of interest that are unavailable and can only be inferred by their similarity with the parameters of other viral illnesses and basic biological processes.

### The model’s demographics

#### Age groups

The basic model considers 4 age groups of healthy hosts: 1) 0 to 12 years of age; 2) 13 to 19 years of age; 3) 20 to 59 years of age; and 4) 60 years of age or older. The healthcare practitioners who work as doctors, nurses and nurse assistants and the healthcare practitioners who work in retirement homes belong to the 20–59-year age range.

#### Living spaces

1) home, where a family is living; 2) working place, where adults meet in working hours; 3) Children’s school, where the hosts in the 0-12 years old range meet in school hours; 4) Teen Agers school, where the hosts in the 13-19 years old range meet in school hours; 5) public spaces, as streets, shops, and public transportation where hosts move from one to other place; 6) Leisure areas, including week-end massive uncontrolled street drinking meetings, involving adolescents and a proportion of adults; 7) Elderly Day Centre, where a 40% of elderly host meet daily; 8) Elderly Nursing Home, where a group of elderly hosts have a permanent stay); 9) Hospital Wards, attended by patients with severe symptoms, or by patients with critical symptoms waiting for a bed in the Intensive Care Unit; 10) Intensive Care Unit, the place in Hospital where critical patients are admitted until reaching the carrying capacity; 11) Post-ICU Setting, where patients that have been discharged from ICU because good prognostic stay for a week before coming back home, although as patients with a length of stay of app. 10 days are unfrequently transmitters (*Kampen et al., 2020*).

#### Population structure at home

We model the following compositions (the number of families is indicated in parenthesis): a) 1 external adult worker, 1 internal adult worker, 2 children less than 12 years old (175 families) ; b) 1 external adult worker, 1 internal adult worker, 1 children less than 12 years old and 1 adolescent (30); c) 1 external adult worker, 1 internal adult worker, 2 adolescents (105); d) 1 Hospital or Elderly Nursing Home worker, 1 adult home worker, 2 small children (30); e) 2 home-working elderly people (1190); f) 1 adult external worker, 1 adult internal worker (496); g) 1 adult external worker, 1 adult internal worker, 1 small child (276); h) 1 adult external worker, 1 adult internal worker, 1 teen ager (184); i) 2 external adult workers, 2 children less than 12 years old (175) ; j) 2 external adult workers 1 children less than 12 years old and 1 adolescent (30); k) 2 external adult workers, 2 adolescents (105); l) 2 external adult workers, 1 small child (276); m) 2 external adult workers, 1 teen ager (184); m) 2 external workers (744).

#### Division of time for the various hosts

The model has the following divisions of time: 1) For external adult workers – Mon-Fri 07:00–08:00 travel in public spaces, 08:00–16:00 working at the workplace, 16:00–17:00 travel in public spaces, and 17:00–07:00 at home. 2) Internal home workers start working at 08:00, 09:00, 10:00, 16:00, 17:00 or 18:00 (with 10% assigned to at each of these times). They have a option of outside activity (e.g., shopping) for 1 (40%), 2 (24%), or 3 h (36%) every day except Saturday and Sunday. 3) For children and adolescents: 08:00–09:00 travel in public spaces and 09:00–16:00 at school. The time at which they return home can range from 17:00 to 18:00 (20%), 18:00 to 19:00 (48%), and 19:00 to 20:00 (32%). From then until 08:00, they stay at home, all day except Saturday and Sunday. 4) For hospital workers, the model distinguishes between day shift workers (07:00–08:00 travel in public spaces, 08:00–16:00 in hospital wards/ICU, 17:00–19:00 travel in public spaces, and 19:00–7:00 at home) and night shift workers (18:00–20:00 travel in public spaces, 20:00–06:00 in hospital wards/ ICU, 06:00–07:00 travel in public spaces, and 07:00–18:00 at home. 5) Care homes are attended by 20%–40% of retired workers over 60 years of age, with the following division of time: 08:00–09:00 travel in public spaces, 09:00–18:00 in the care home, 18:00–19:00 travel in public spaces, and 19:00–08:00 at home. 6) Retired older adults not living in care homes have a division of time identical to that of internal home workers. 7) In care homes for the elderly, elderly people live together permanently within a common space, with 5 adult external healthcare workers having a division of time similar to that of hospital healthcare workers. For the divisions n hours, note that we are considering that epidemics occur in the winter-spring, but these data can be adapted to the summer-autumn if necessary.

The model considers other activities. External and internal workers, children and adolescents might go out to enjoy public spaces during the weekend, starting a 10:00 (30%), 17:00 (35%) or 18:00 (32.5%) for 1 (40%), 2 (24%) or 3 h (36%). During the dawn and early morning and on Friday nights, a high proportion of adolescents (50%) and a small proportion of adults (15%) are located in leisure areas. On Saturday night, the relative proportions of adolescents and adults are 80% and 30%, respectively. These groups are in public spaces from 0:00 to 1:00, in leisure areas from 01:00 to 06:00, and once again in public spaces from 06:00 to 07:00. Forty families (with 4 members) have one or more relatives in care homes for the elderly; each of these family members has a 25% probability of visiting the care home on Saturday and a 25% probability on Sunday. These members are therefore in public spaces from 09:00 to 10:00, in the care home from 10:00 to 16:00 and back in the public spaces on their way home from 16:00 to 17:00.

#### Population numbers

In the model, we consider a representative original sample, consisting of an isolated community (we do not consider the introduction of foreigners, but this will be presented in a forthcoming study) of 10,320 healthy individuals (a number that reflects the demographic adjustments and limitations of the computer workload), consisting of 1372 children attending school, 848 adolescents attending school, 4324 external workers, 1266 adult home workers, 2380 elderly home workers, 20 hospital-based professionals (5 in wards and 5 in the ICU, on day and night shifts), 100 elderly individuals living permanently in nursing homes, served by 10 adult external health workers (5 per shift). Overall, we have 1372 young children, 848 adolescents, 5620 hosts aged 20 to 59 years, and 2480 hosts aged more than 59 years.

#### System rules on the population dynamics of infected hosts

1) Asymptomatic hosts do not modify their habits. 2) When symptomatic patients perceive their first symptoms, they might have a maximal viral load of approximately 70% (see following paragraph). They might initially consider the infection as mild and remain at home until their clinical cure (again asymptomatic), for an average of 7 days since the first symptoms. 3) A host in an elderly nursing home with mild infection remains in the nursing home. 4) For each hour that elapses, there is a 0.0015 probability that the host with mild infection progresses to severe infection, in which case there are admitted to a hospital. Therefore, the possibility that a host with mild infection who has not recovered after the first week will progress to a severe infection is 22.29% after 1 week and 39.61% after 2 weeks. 5) Patients with severe infection have a mortality risk of 0.0005/h, 8.05% the first week and 15.7% the second week; 6) Patients with severe infection admitted to a hospital become critical at a rate of 0.002/h, which is 28.56% in 1 week and 48.97% in 2 weeks. In this case, the patients are admitted to the ICU or remaining at the hospital waiting for ICU admission. 7) Critical patients have a mortality risk of 0.002 per hour in the ICU (28.56 in 1 week and 48.97 in 2 weeks) and 0.004 per hour if they could not be admitted to the ICU (49% in 1 week and 73.99 in 2 weeks). 8) Patients who are cured in the hospital (with either severe or critical infection) stay an average of 1 week longer in the hospital, after which they are discharged home.

### Conceptual and parametric framework of the basic model

#### Viral load

Viral load refers to the number of infectious viral particles in the respiratory secretions of an individual who has interacted with the virus. This number should correlate with the tissue culture infectious dose (TCD50), the quantity of viral particles able to infect 50% of inoculated tissue cultures (*Bordia et al., 2020*), which can be estimated at approximately 400 particles/mL. *In vivo*, this number is highly variable and difficult to measure but has been estimated at 10E+7 per mL of respiratory secretions (*Pujadas et al., 2020)*. In this study, we considered the viral load in an individual as a fraction of the maximal viral load. For instance, a viral load of 0.001 means that the load is a thousand times lower than the maximal viral load. Infected children do not differ from adults in terms of viral load (*Colson et al., 2020*). Patients with bronchopulmonary damage or pneumonia have remarkably high viral loads. In reverse transcription polymerase chain reaction (RT-PCR) assays, the cycle threshold (*Ct*) is based on the number of cycles required for the fluorescent signal to exceed the background level and can be inferred as a surrogate for viral load. If the *Ct* is more than 24 cycles (fewer than 20,000 viral copies/mL), the tested individual has a significant reduction in their ability to transfer the virus, and the transfer does not occur with a *Ct* >30-34 (*La Scola et al., 2020*). However, this does not mean that these patients (particularly ones with severe infection) do not carry viable viruses (*Singanayagam et al, 2020*). Of course, protective measures (e.g., masks) reduce the acquiring and spreading of viral loads and consequently transmission.

#### Mode of transmission

The virus is transmitted to another individual through contact with the respiratory secretions of an infected individual (with or without symptoms), directly reaching the respiratory tract or indirectly through hand-respiratory tract (nose, mouth) contact. There should be a “minimal propagulum” for viral loads, a threshold below which no significant (permanent) transmission takes place (estimated at fewer than 20,000 copies/mL) (*Bullard et al., 2020*). When crossing this threshold, there is a certain probability for contagion, the rate at which an infected patient can transmit the virus to non-infected individuals. This rate is influenced by the donor’s viral load. Given that the samples are frequently heterogeneous, the viral load should be determined as the delta *Ct* (the difference between the viral target *Ct* and a *Ct* marker of human cells) but more importantly by the transmission mode (rhinorrhoea, sneezing, frequent coughing, and, in particular, sputum formation, which produce large airborne droplets, multiplying the risk of transmission by 4–5-fold), thereby correlating with clinical severity (*Guan et al. 2020*), previous respiratory disease, age, and habits. For instance, children can have remarkably high viral loads with low contagion efficiency. These factors could explain the phenomenon of so-called “superspreaders”. There are apparently numerous differences in viral loads when comparing asymptomatic and non-severe symptomatic patients (*Lee et al., 2020*).

#### Infectious period

This period refers to the duration of infectiveness, the time duration which a person harbouring the virus is able to transmit that infection to another human. In our model, we consider that on average a colonized individual can transmit viruses 48 h before symptom onset. In a given individual, the maximal transmission rate occurs from the second to fourth day after symptom onset. In previously healthy patients (children and adults), the danger of transmission is minimal 1 day after symptom onset. In patients with bronchopulmonary damage or pneumonia, the infectious period is longer (15–20 days).

#### The contagion index

Infected hosts who exceed a viral density threshold have a certain possibility of infecting an infection-naïve host, a possibility that obviously depends on the contact rate, which refers to the number of non-infected individuals a person harbouring the virus can be expected to meet. This number depends on numerous factors, including population density, number of contacts within 100 cm/h, amount of time spent in closed areas, and climatic factors. Entering parameters reflecting these factors into the model is difficult, given the heterogeneity and highly variable nature of these situations. Existing models of COVID-19 contagion rely on parameters such as the basic reproduction number (*R0*), which represents the number of secondary infections originating from a primary infected individual in a fully susceptible population and is difficult to measure in real time (which is one of our objectives with this model). These models employ static statistical methods that do not capture all of the relevant dynamics (*Oehmke et al., 2020*).

In our model, we dissect the contagion dynamics by considering the population dynamics of the infected hosts according to age, work and leisure activities, living spaces and public spaces, which requires the consideration of an estimated ***contagion index per person/h***. For instance, a basic contagion index of 0.125/h for hosts with a viral load of over 20,000 copies/mL (0.2% of the maximal virus load, 10E+7/mL) means that in a given setting an infected host in contact with other hosts for 8 h produces on average 1 contagion (8 h × 0.125/h). Lower contagion indexes (effective daily contagions per host) have been proposed (*Casares and Khan, 2020)* but have proven to be unrealistic in preliminary test runs of our model (data not shown). To mimic the contagion window, our model includes a specific time-counter for each type of host to adjust for the time when the host starts to spread the virus.

#### Natural immune response and receptivity to contagion

Some of the individuals exposed to the virus do not develop symptoms, even when lacking acquired immunity. This lack of symptoms could be due to the rapid elimination of most if not all arriving viral particles by a highly efficient innate immunity (mostly mediated by interferons and resident macrophages, see below) (*Dogra et al., 2020*), eventually eliminating the propagule, particularly in children and young adults, thereby having a significant effect on contagion.

Contagion is highly influenced by the infected host’s immune response. The correlation between antibody-mediated immunity to the coronavirus and protection and severity has been documented (*Huang et al., 2020*) but less documentation exists for the natural (innate) immune response. In general, there is scarce information on the relationship between the immunological status and the transmission rate. For the purposes of this model (which does not necessarily fully reflect reality), we differentiated four types of host populations accordingly to their natural immune response.

First, there are those who acquired the virus, which can replicate until reaching 20% of the maximal viral load. During this process, the innate immunity is triggered, causing the viral load to decrease. An acquired immune response does not occur. These cases are either asymptomatic or have very mild symptoms. This type is here designed as the “efficient innate immunity/lacking acquired immunity/mild to no symptoms” (E-inn/L-acq/N) type. Second, there are those in whom the viruses are efficiently cleared by their innate immunity but that cross the viral load threshold for triggering acquired immunity. In most cases, these hosts remain asymptomatic or have mild symptoms. This type is designed as the “efficient innate immunity/normal acquired immunity/mild to no symptoms” (E-inn/N-acq/N) type. Third, there are the hosts whose innate immunity is insufficient for reducing the viral load, which increases accordingly and crosses the threshold after which symptomatic infection occurs and acquired immunity is developed. This type is designed as the “inefficient innate immunity/normal acquired immunity/Symptomatic” (I-inn/N-acq/S) type. Fourth, there are the hosts whose innate immunity is insufficient to clear the virus, resulting in a symptomatic infection; however, their acquired immune response is weak or slow. This type is designed here as the “inefficient innate immunity/weak acquired immunity/Symptomatic” (I-inn/W-acq/S) type. The E-inn/L-acq/N and I-inn/W-acq/S types can be reinfected after exposure with a contagious host but will respond to the new challenge according to their specific response pattern. Table 1 lists the estimated frequency of these populations per age group that were used in our model.

**Table 1:** Estimated frequency of immunological response types per age group.

|  | Age groups, years |  |  |  |
| --- | --- | --- | --- | --- |
|  | 0–12 | 12–19 | 20–60 | >60 |
| E-inn/L-acq/N | 0.1979 | 0.1956 | 0.12 | 0.1562 |
| E-inn/N-acq/N | 0.7421 | 0.6844 | 0.52 | 0.2437 |
| I-inn/N-acq/S | 0.03 | 0.06 | 0.18 | 0.3 |
| I-inn/W-acq/S | 0.03 | 0.06 | 0.18 | 0.3 |

#### Contagions per age, severity, and location

In practice, contagions essentially occur when hosts of the I-inn/N-acq/S and I-inn/W-acq/S types interact with infection-naïve hosts. Non-symptomatic hosts are low transmitters, in general those who, after being exposed to the virus, were able to clear the infection by innate immunity and are expected to have a low or non-existent viral load. We considered that these hosts have an average transmission rate of <0.01 per person/h. Non-symptomatic hosts that progress to a more severe status have a transmission rate of 0.02 person/h. Table 2 presents the estimated contagion index per contact person and hour, considering the spaces where the hosts are located, their age and the severity of the infection.

**Table 2.** Estimated contagion index per contact person and hour, considering the spaces where the hosts are located, and the age, immunological and clinical groups to which they belong. The “x” symbol represents impossible situations (e.g., individuals with severe or critical symptoms cannot attend their workplace).

|  | **Public spaces** | **Homes** | **Workplaces** | **Elementary schools** | **Secondary schools** | **Leisure locations** | **Nursing homes** | **Hospitals** |
| --- | --- | --- | --- | --- | --- | --- | --- | --- |
| **E-inn/N-acq/N or E-inn/L-acq/N**<br>**Asymptomatic** | 0.02 | 0.02 | 0.02 | 0.03 | 0.03 | 0.06 | 0.04 | 0.05 |
| **I-inn/W-acq/S or I-inn/N-acq/S**<br>**Incubation for disease** | 0.1 | 0.1 | 0.1 | 0.15 | 0.15 | 0.3 | 0.2 | 0.25 |
| **I-inn/W-acq/S or I-inn/N-acq/S**<br>**Weak symptoms (0–60 years)** | 0.1 | 0.1 | 0.1 | 0.15 | 0.15 | 0.3 | 0.2 | 0.25 |
| **I-inn/W-acq/S or I-inn/N-acq/S**<br>**Weak symptoms (>60 years)** | 0.2 | 0.2 | x | x | x | x | 0.4 | 0.5 |
| **I-inn/N-acq/S**<br>**Severe or critical symptoms** | x | x | x | x | x | x | 0.4 | 0.3 |

#### Reinfections

In our model, a particular infected patient who progressed to a healthy stage reassumed their daily routine, where they might interact with other infected hosts, exposing the patient to a potential new viral contagion, which might progress to a new infection. Reinfection of a cured patient depends on their immunological status. If they are an E-inn/N-acq/N or I-inn/N-acq/S type, reinfection does not occur. If they belong to the E-inn/L-acq/N type, the second viral challenge will be aborted because it occurred in their first infection. Acquired immunity is in fact critical for preventing reinfection (*Kim et al., 2021*). Accordingly, if the recovered and now healthy patient were of the I-inn/W-acq/S type, the probability of reinfection in the model depends on the intensity of the immune response, which can be weak (such as the during the first contagion) or more effective than during the first contact. We consider both probabilities equivalent (50%-50%).

#### Elderly lockdown

The “elderly lockdown” intervention involves cancelling all external visits to nursing homes, closing elderly day centres, and reducing by 80% the external activities of the elderly.

### Representation of the temporal progression of the epidemic load

#### Frequency of infected hosts

Only a fraction of individuals who acquire the virus are considered “cases”; the rest are able to clear the first viral infection through their innate immunity. We represent the progression of the total number of cases by age group but differentiate those who are infected but remain asymptomatic or have short mild symptoms thanks to the innate immunity from those who are clearly symptomatic but clear the infection through their acquired immunity. We include representations of the frequency of infected hosts with various levels of severity who are admitted to hospitals or ICUs.

#### Frequency of susceptible or immune hosts

Susceptible hosts are those who have not been exposed to the virus and lack sufficient innate or acquired immunity to clear the virus infection. Immune hosts are those who have cleared the virus and have recovered from the infection, either through innate or acquired immunity. A small percentage of individuals who are cured by their innate immunity could become susceptible.

#### Frequency of contagions

We represent the absolute number of contagions in the various types of hosts, behaviours, and compartments. In our model, a contagion is a “rule” applied with a certain probability when each of computational entities representing the various types of infected hosts comes into contact with another host, leading to the acquisition of the virus. The frequency of contagions does not exclude the virus transmission events to already infected hosts. The interest in estimating the frequency of contagions lies in determining the general evolution of the risks associated with particular types of hosts, behaviours or compartments. The frequency of contagion is represented globally but also differentiates the contagions originating from cases that progress to healthy due to innate or acquired immunity.

#### Mortality rate

This representation illustrates the cumulative number of deaths caused by the COVID-19 infection over time, according to age. In the text, we also express the global number as the mortality rate (deaths per 1000 inhabitants per year).

## RESULTS

### Influence of time until the implementation of interventions of varying intensity on the number of the individuals in the COVID-19 response groups

Figure 1 presents the basic epidemiological dynamics of the modelled epidemics. The epidemic starts with the spread of asymptomatic cases that have an efficient innate immunity but have a viral load sufficient for triggering an acquired immunity response. These cases reach a prevalence of 29.07 at 42 days (1000 h) after the onset of the epidemic but decreases to less than 2.5 at 84 days (pink line in Figure 1). This initial pulling wave is followed by the contamination of asymptomatic individuals (mostly children and adolescents) who aborted the infection due to a powerful innate immunity without developing protective acquired immunity. Combined with re-exposures to the virus, the curve drops slowly with time but with no significant transmission to other populations. Symptomatic patients, with insufficient innate immunity to prevent high viral loads, reach maximum prevalence at approximately 4.0–5.0 at 52 days from the onset of the epidemic. In these symptomatic patients, the number able to develop a normal acquired immunity (green line) decreases more rapidly but not excessively compared with those with weak immunity (mostly elderly patients). A residual number of symptomatic individuals are maintained even at 200 days from the onset of the epidemic.

**Figure 1.**
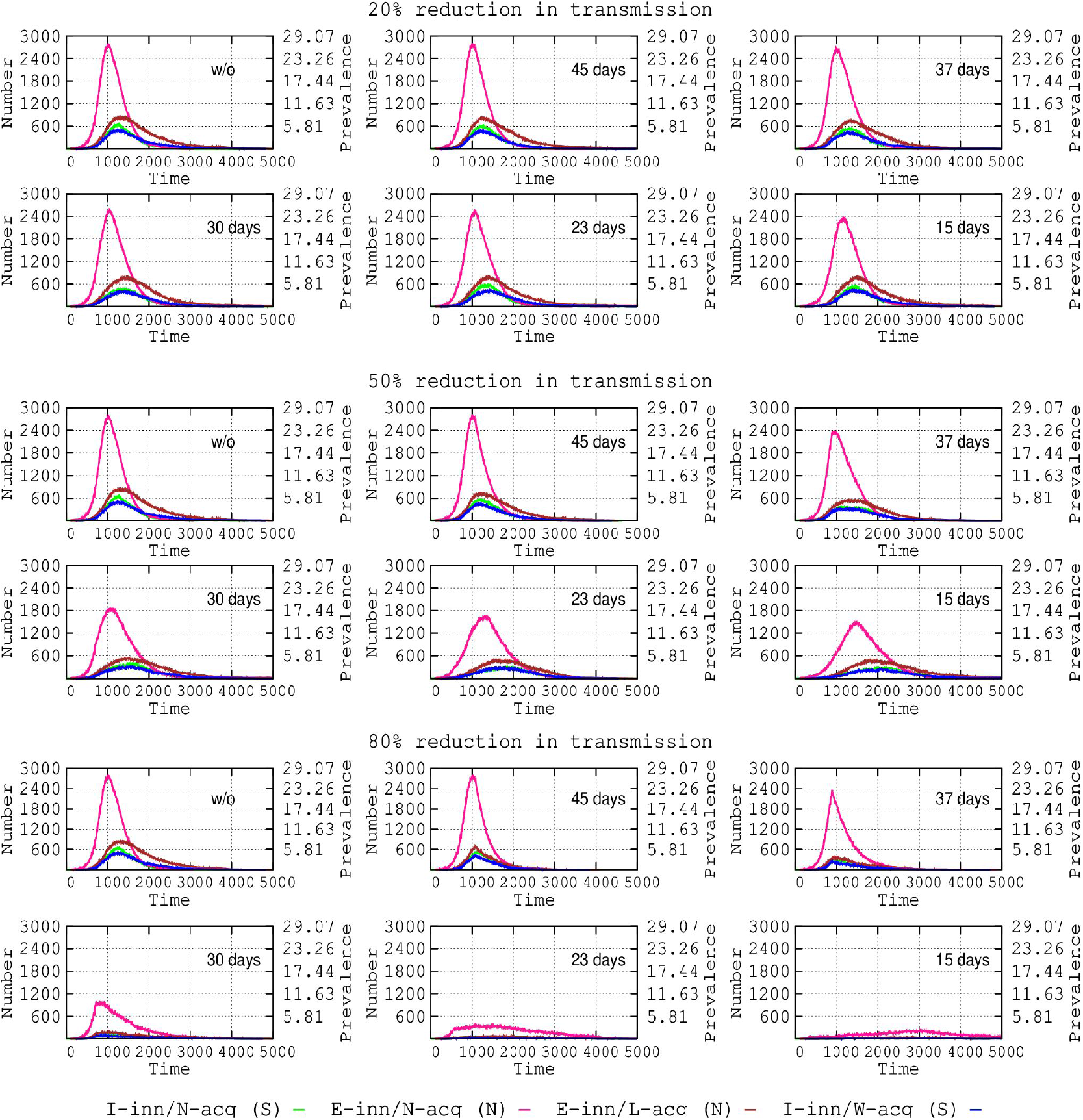
Influence of interventions (time of adoption and intensity of reduction in transmission) on the number of individuals in the host populations with varying response to the COVID-19 infection. I-inn/N-acq/S (symptomatic): insufficient innate immunity, normal acquired immunity (green line); E-inn/N-acq/N (non-symptomatic): efficient innate immunity, with normal acquired immunity (pink); E-inn/L-acq/N (non-symptomatic): efficient innate immunity, lacking or with very weak acquired immunity due to poor antigenic challenge, violet; I-inn/N-acq (S, symptomatic): insufficient innate immunity, weak acquired immunity (dark blue). Steps in the time scale represent hours after the onset of the epidemics (approximately 1000 h, 42 days, approximately 5000 h, 7 months).

### Reduction in the maximum prevalence in the COVID-19 response groups: influence of time until implementation of interventions of varying intensities

In this section, we model the effect of time from the start of the epidemic to the application of a counteractive intervention. We expect these interventions to reduce the disease contagion by 20%, 50% or 80%. The effects on four populations with differing immunological responses are presented in Figure 2.

**Figure 2.**
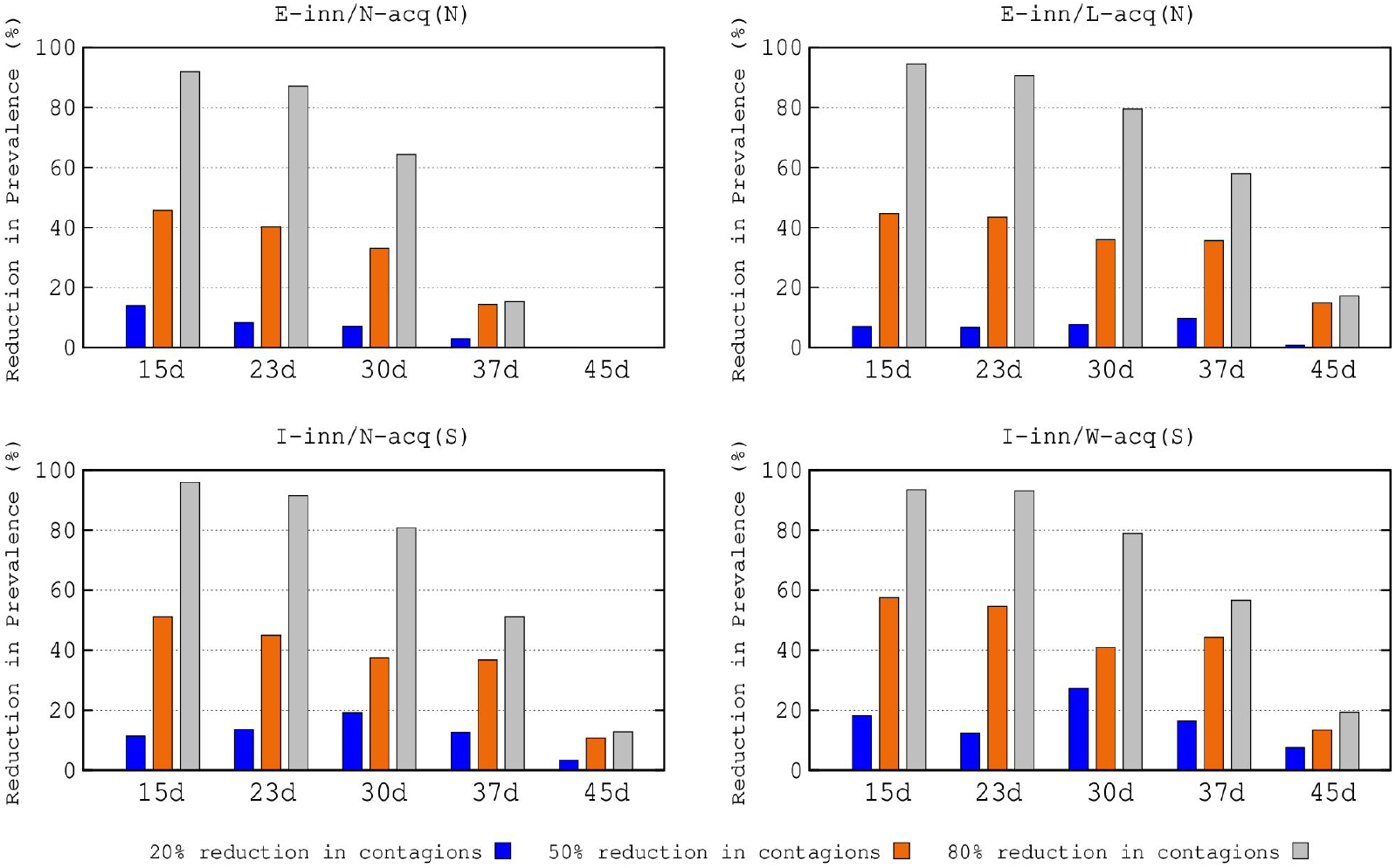
Percentage reduction in maximum prevalence (RMP) in the various COVID-19 immunological response populations (see text of Figure 1) according to the time interventions were adopted and according to the intensity of the reduction in transmission.

The early implementation of interventions that can decrease transmission 15 days after the start of the epidemic reduces the maximum prevalence percentage for all groups. However, if the intensity of the reduction is low (20%), the reduction in the maximum prevalence is modest: 7%–13% for non-symptomatic cases (E-inn/N-acq and E-inn/L-acq) and 10%–18% for symptomatic cases (I-inn/N-acq and I-inn/W-acq). With a medium intensity of reduction in transmission (50%), the reduction in the maximum prevalence is 42%–45% for non-symptomatic cases and 50%–58% for symptomatic cases. Combining the early implementation of interventions and high intensity, 80% of the reduction in transmission, the maximum prevalence values strongly decrease below 90% (90%–95% for non-symptomatic cases and 92%–95% for symptomatic cases).

The differences between the initial implementations to decrease transmission at 15 or 23 days, for any level of intensity, are relatively minor. If these actions are promoted after 30 days, the effects in reducing maximal prevalence are maintained and remain modest at 20% intensity. At a 50% intensity of reduction, we achieve lower reductions in maximum prevalence after 30 days in transmission for non-symptomatic cases (32%– 35%) and symptomatic cases (38%–40%). Even at an 80% intensity of reduction, there is a clear drop in the reduction of maximum prevalence (65%–79% for asymptomatic cases and 79%–81% for symptomatic cases). If interventions are adopted at 37 days after the onset of the epidemic, the reductions in maximum prevalence rates are relatively modest compared with these obtained at 30 days for transmission reductions of 20% and 50%. However, even with an intensity of 80% in transmission reduction, a strong decline in maximum prevalence occurs for symptomatic patients (from 79%–81% at 30 days to 52%–57% at 37 days). Interventions adopted at 45 days from the onset have an extremely limited effect in reduced the maximum prevalence for the various groups. The numerically important E-inn/N-acq/S group has no reduction at all. Even with a high intensity in reducing transmission (80%), the prevalence of symptomatic cases (E-inn/N-acq and E-inn/W-acq) was reduced by only 12% and 19%, respectively.

### Influence of time until the implementation of interventions of varying intensity on the populations at given clinical stages

Figure 3 (upper row) represents the influence on the populations in different clinical stages of the interventions for reducing transmission with varying intensities (20%, 50%, 80%) and adopted at different time periods from the onset of the epidemic. In general, the early adoption of interventions to decrease transmission increases the **reduction of the maximum prevalence** (RMP, represented in ordinates) for all populations. RMP also steadily increased due to more intense transmission reductions. An apparently paradoxical trend is apparent when a 20% reduction in transmission (blue bars) increases the RMP to a higher degree in the patients progressing to higher severity. Conversely, an 80% reduction (grey bars) increases the RMP slightly more in asymptomatic and weak symptomatic patients. In general, no relevant RMP differences are found if the implementation of transmission reduction occurs after 15 or 23 days after the onset of the epidemic, but the RMP decreases significantly with longer delays. The detailed values are represented in Table X in the Supplementary material.

**Figure 3.**
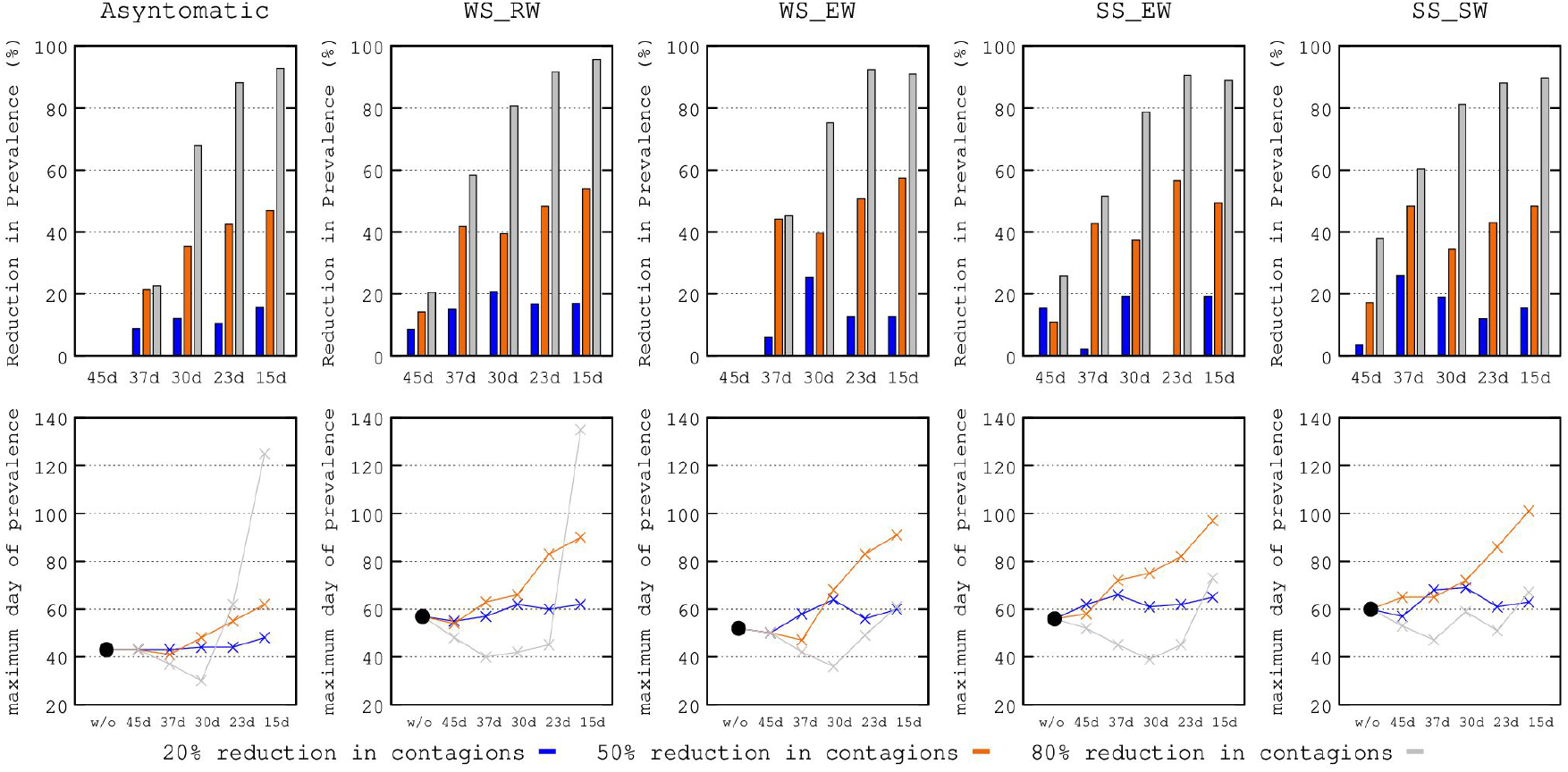
Upper row: **percentage reduction in maximum prevalence** (RMP, in the number of cases) according to the time of adoption and intensity of interventions (20% blue; 50% red; 80% grey) in the various clinical progression groups: asymptomatic, WS-RW (weak symptomatic rarely progressing to worsening), WS-EW (weak symptomatic progressing to worsening), SS-EW (symptomatic progressing to worsening), and SS-SW (symptomatic progressing to severe or critical worsening). Lower row: **delay in maximal prevalence** (DMP, in days) of the various clinical progression groups. Black dots represent the days from the onset of the epidemic where the maximum prevalence occurs in the event of no-intervention.

Figure 3 (lower row) shows the influence of the implementation time and the intensity of interventions for reducing transmission on the **delay in maximum prevalence** (DMP, expressed in days) of the populations at different clinical stages. The black dots represent the expected DMP if no interventions were adopted. There is a sequential process, where infected but asymptomatic patients can progress to WS-RW or WS-EW, WS-EW can progress to SS-EW and these can progress to SS-SW (see definitions in the legend of Figure 3). The DMPs therefore steadily increase over time in balance with the severity and the maximum prevalence of the more severely ill patients occurring at later stages. Therefore, the early adoption of interventions (e.g., 15 days after the onset of the epidemic) ensures that the effect is extended in time, influencing all stages in which patients potentially worsen. This explains why an 80% reduction in transmission (grey line) adopted at 15 days pushes the maximum prevalence peak of asymptomatic patients from less than 50 days in the case of non-intervention to more than 100 days. However, the same 80% reduction adopted at 45 days has much less effect on symptomatic patients, because this late intervention has arrived too late to decrease the number of asymptomatic patients and thus their potential progression to severity. Moreover, as the emergence of severely ill patients is naturally delayed, the late adoption of interventions results in insufficient time to significantly act on the process. An intervention with only a 50% reduction in transmission (red line) adopted before 30 days results in a clear delay in maximum prevalence in all stages of severity, including the more severely ill patients.

Figure 4, represents (on an exponential scale) the influence of adoption times of interventions of varying intensity for preventing transmission on the **extent of the time period** (ETP) where the different clinical severity groups are maintained at different prevalences. A decrease in asymptomatic cases is associated with a longer maintenance (purple and green lines), particularly if the transmission-reducing interventions are implemented very early (a comparison of the adoption of interventions at 37 and 15 days is shown in the red boxes of Figure 4). In general, the early adoption of transmission-reducing interventions reduces the prevalence; however, the emergence of new cases is extended in time and applies to symptomatic and critical patients (compare the blue and black lines in the red boxes), which should influence the prolongation of the epidemic, likely by maintaining the sources of contagion for successive waves (not considered in this manuscript) while favouring the dynamics of hospital and ICU admission by reducing the risk of exceeding bed capacity.

**Figure 4.**
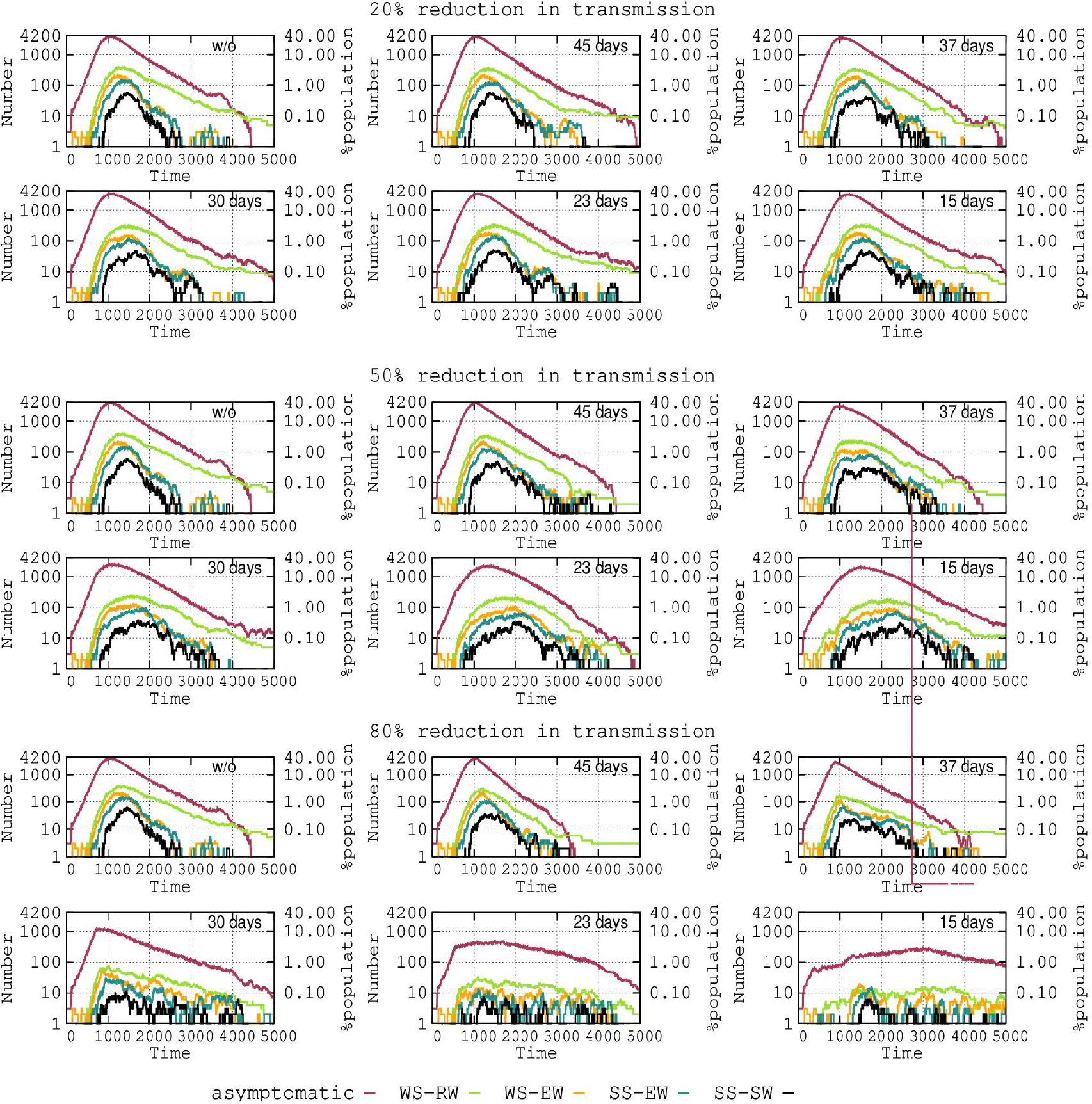
Influence of adoption times for interventions of varying intensity on the extent of the time period (ETP) where the number of individuals belonging to the different clinical severity groups are maintained.

### Influence of time until the implementation of interventions of differing intensities on the mortality rate per age group

We estimated the age-related mortality in the absence of interventions in our model as follows: <0.1% for the 0–12-year group, 0.29% for the 13–19-year group, 1.64% for the 20–59-year group and 17.58% for >60-year group. Figure 5 shows the progression of mortality for these age groups when varying intensities (%) of transmission reduction are adopted at different times. The curves corresponding to the 0–12 and 13–19-year groups are not noticeable given the very low mortality rate. The mortality for the 20–59-year group is scarcely influenced by the 20% and 50% reductions in transmission and by the 80% reduction only if it occurs during the first 30 days after the onset of the epidemic. As expected, most deaths occurred in the elderly (older than 60 years, green line). In general, the mortality rates did not increase significantly after 3000 steps (approximately 125 days after the onset). An 80% reduction in transmission significantly decreases the mortality in the elderly patients, even if the intervention is applied at 45 or 37 days after the onset of the epidemic; if adopted earlier (e.g., 23 or 15 days), deaths can be almost completely prevented.

**Figure 5.**
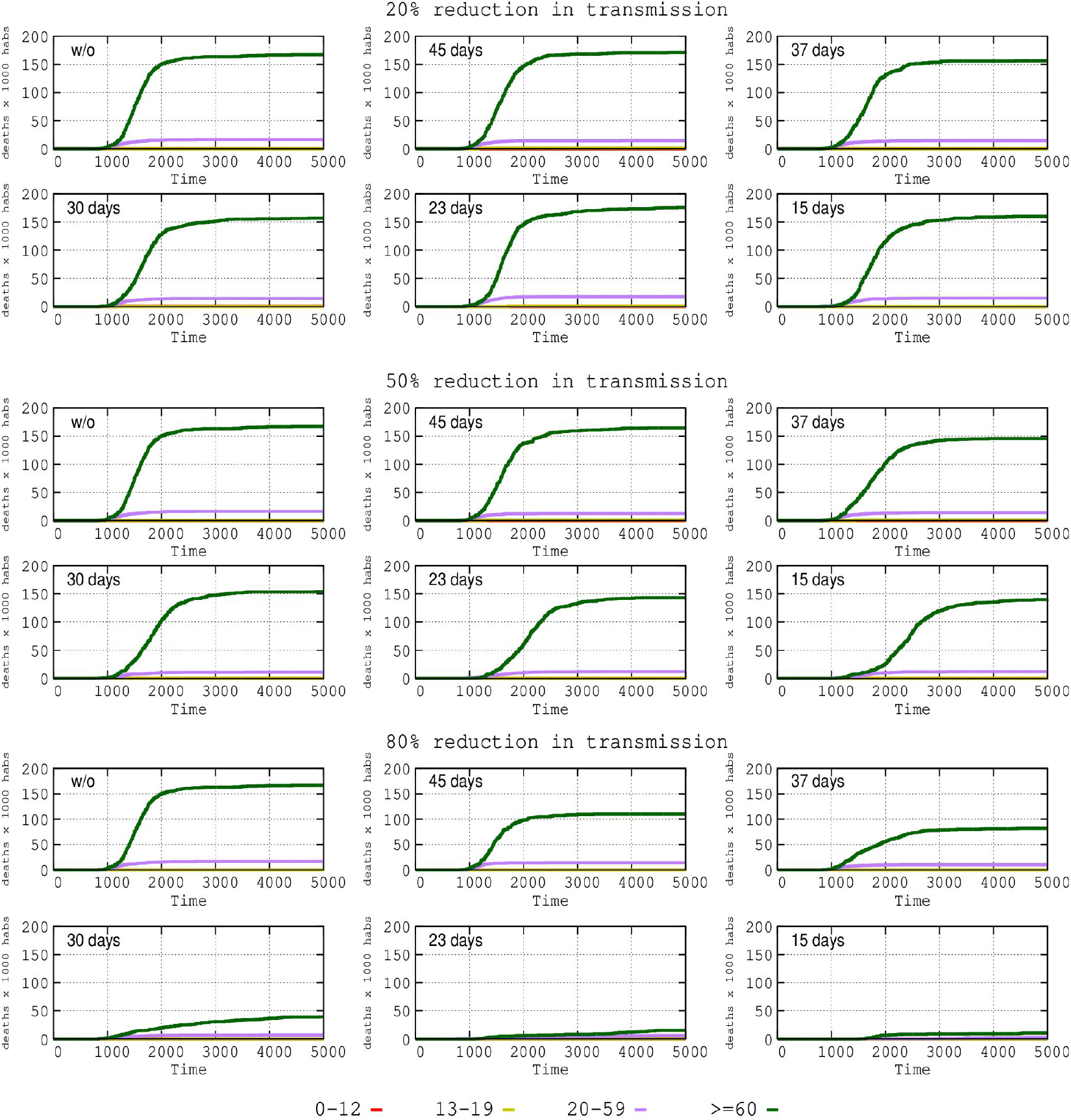
Influence of time until the implementation of interventions of varying intensity on the mortality rate per age group: 0–12 years (pink), 13–19 years (red), 20–59 years (violet) and > 60 years (green). In this representation, the lower age groups are not visible due to the low mortality rate.

### Elderly lockdown

Considering the above results, we tested the effect of an “elderly lockdown” in the absence of other general interventions. The “elderly lockdown” intervention implies suspending all external visits to nursing homes, the closure of elderly day centres, and an 80% reduction in external activities for the elderly (>60 years). In nursing homes and hospitals, we tested the effect of supplementary measures for reducing transmission by 80%, 90% and 95%.

We first modelled the effect of this elderly lockdown intervention in the absence of any other intervention (including general measures to reduce transmission in the general population) on the general dynamics of the epidemic. As shown in Figure 6 (top row), the intervention reduced the prevalence in non-symptomatic cases by approximately 25%– 30% in the general population, and the reduction of transmission, even of 95%, in elderly areas add very few extra reductions in prevalence.

**Figure 6.**
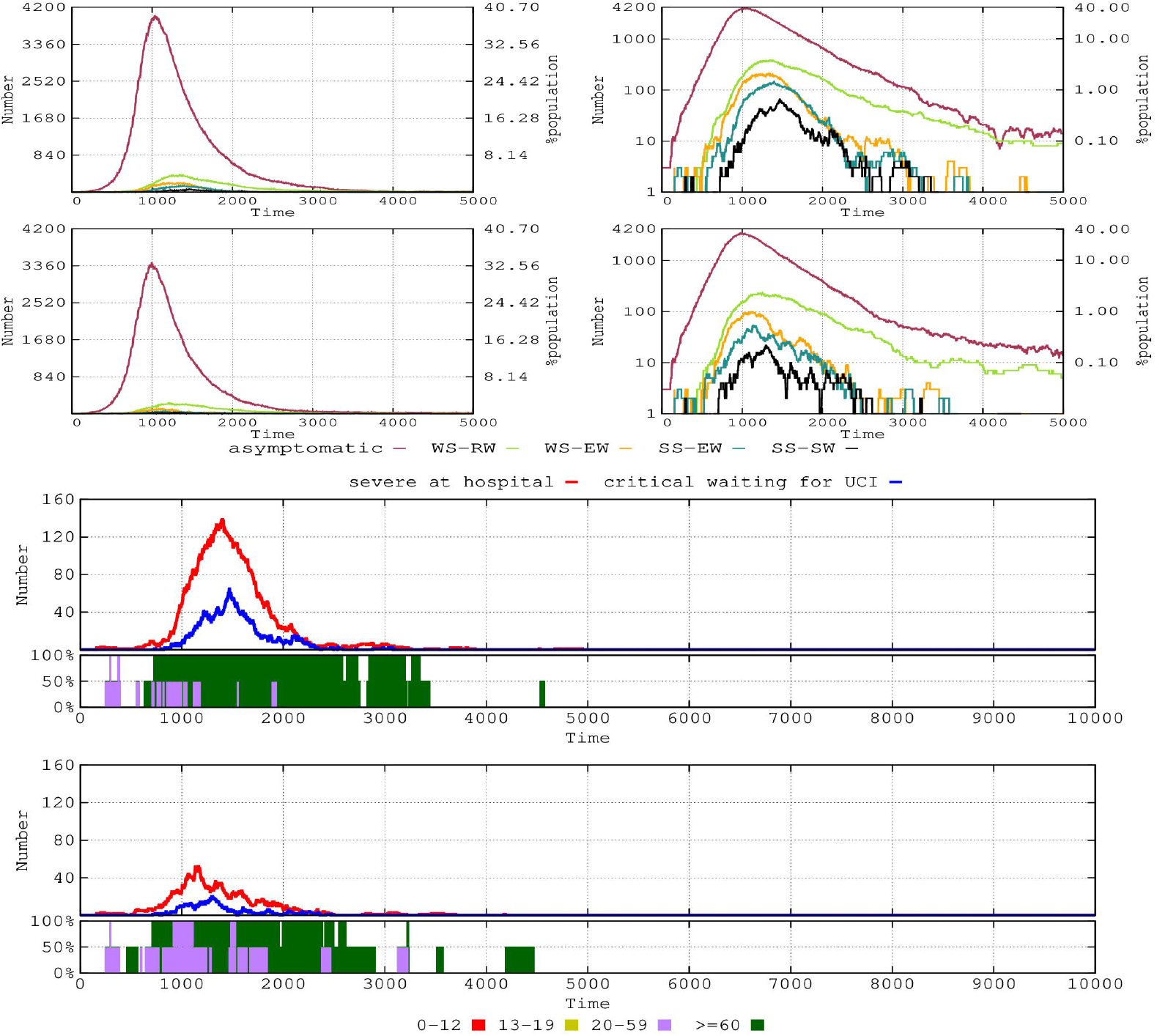
Effect of the elderly lockdown (older than 60 years) in the absence of any other general intervention on the number of individuals of the various immunity response groups (upper two rows, logarithmic scale at the right). In the lower two figures, hospitalised severely ill (red line) and critical (blue line) patients; in the boxes below, the percentage of hospital ICU use by patients aged 20–59 years (violet) and older than 60 years (green).

The effect of the lockdown on the prevalence of infection in the elderly population (Figure 6, upper and lower panels) is certainly impressive. The prevalence of infection dropped by 90% in the non-symptomatic cases and 85%–90% in the elderly symptomatic cases. The extra addition of transmission reducing interventions by 95% has a negligible effect in further drops in prevalence but shorten the time where symptomatic cases occur. The elderly lockdown also had a major effect on the availability of ICU beds (lower panel of Figure 6). Without restrictions, the ICU is fully occupied most of the time, and, except at the start of the epidemic (approximately 40 days, 1000 hourly steps in Figure 6), where a substantial number of severely ill patients in the 20–59-year age range are admitted (violet bars), the elderly patients (green bars) were predominant for an extended period (at least 6 months), and even scattered cases occurred later. The number of cases in the hospital was high (red line), and many critical cases deserving intensive care were waiting for ICU admission (blue line). With the elderly lockdown as a single intervention, more people in the 20–59-year age range can be treated, with much less pressure from elderly patients and for a shorter period.

## DISCUSSION

Epidemic processes occur within complex and stratified landscapes, where the possibility of quantitatively ascertaining the different combinations of factors is frequently far beyond the technical capabilities. We present here two examples. The first, which is particularly relevant for the epidemiology of COVID-19, is how to measure the number of individuals who have been exposed to the virus without developing any symptoms of the infection (or very mild symptoms) and do not develop any detectable humoral response. Such a visible response to exposure relates to the innate protective response, which is understood here as the ensemble of factors preventing the virus from producing an infection, from innate immunity to the scarcity of suitable viral receptors in the cell, (*Cox and Brokstad, 2020*), or perhaps the interference of the virus by *Bifidobacterium*, frequent in children and young people (*Ali, et al., 2021)*. In those patients who have been infected but do not develop illness, the viruses probably have a lower transmission rate to susceptible individuals; however, this cannot be measured. A second case in point is our inability to predict the epidemic dynamics in a susceptible human population when the first COVID-19 cases emerge, when various measures are adopted within different time frames. How do we ascertain the effect of a single intervention? How do we to establish an identical control population where the intervention has not been implemented? Modelling is the only way to approach a rational management of epidemic situations (*Sridhar and Majunder 2020; Gibney, 2020*).

Membrane-computing, derived from natural and cellular computing, creates a virtual community of hosts, a community of computable objects in the model, composed of immunologically different individuals of varying ages and social activities, able to stochastically interact at particular times and to react differentially with the infective objects, i.e., the virus (see Introduction). The model is in fact a fine-grain recreation or virtual representation of reality. With our agent-based model, we attempted to meet the challenge posed by Kathleen O’Reilly at the London School of Hygiene and Tropical Medicine: “You need to collect information on households, how individuals travel to work and what they do at the weekend” (*Adam, 2020*). Of course, the interactive parameters are unknown or known imprecisely, but they can be established as ranges obtained from a meta-analysis (*Fonfría et al., 2020*).

This study’s consideration of the effect of transmission interventions on various populations with varying immunologically associated responses to viral exposure, the different associated risks of virus transmission, and the progression of the clinical prognosis, occasionally leading to death, was scarcely explored in previous modelling approaches. The definition of these populations and their risks was based on a relatively scarce amount of objective data and implied certain simplifications. For instance, young children have an immature immune system and are frequently infected by viral organisms; however, COVID-19 symptomatic cases in children are uncommon (*Kloc et al., 2020; Calvo et al., 2020, O’Driscoll et al., 2021)*. As stated above, we attribute this fact to an “effective innate immunity”, understanding as effective innate protection the ensemble of factors that prevents COVID-19 growth and infection, unrelated to the immune acquired response (see above). In fact, children acquire COVID-19 from adults, but the transmission rate from children to adults is low, perhaps due to low viral loads in asymptomatic children, combined with weak, infrequent coughing (*Lee and Raszka, 2020; Posfay-Barbe et al, 2020; Fretheim, 2020*). We also included in the model the reasonable assumptions that high viral load correlates with transmission, severity, and mortality (*Van Damme et al, 2021, Pujadas et al, 2021*). Older ages are related to higher viral loads and viral shedding (*To et al. 2020*).

Our results on non-pharmaceutical interventions (essentially targeting transmission) complement the novel aspects previously published in observational studies and in the mathematical modelling of COVID-19 epidemics, including classical (SIR -Susceptible-Infected-Recovered) and others, such as GitHub tools that analyse epidemiological scenarios (*Flaxman et al., 2020; Bhatt 2020; Stokes et al., 2020*). Our computational model mimics the dynamics of a single community with a limited number or inhabitants (approximately 10,000) and isolated from other sources of infection. This precludes us from predicting the “global dynamics” of the infection and makes it difficult to compare some of our results with other modelling estimates, but in general the differences are weak. For instance, our predicted “mortality peak” in the absence of interventions occurs earlier (a little more than 2 months) than in other model estimates (*Ferguson et al., 2020*). However, the model allows for a realistic prediction of the consequences of applying interventions.

The main results of this study (many other results can be obtained with the same model by varying the parameters) include the key influence of the adoption time of transmission-reducing interventions (*Poland, 2020*). The earlier, the better; however, our model showed no clear differences when adopting interventions at 15 or 21 days from the onset of the epidemic. Interventions adopted at 37 days and particularly after 45 days have, however, a weak effect, except in reducing the more severe cases in the elderly groups, particularly with a transmission reduction of 80%. If this strong reduction is applied at 15 or 23 days, deaths can be prevented almost totally. Our detailed analysis of the clinical progression groups indicates that this occurs due to an early intervention reducing the number of individuals who might progress to worse outcomes, including asymptomatic or mild symptomatic patients. Interestingly, the early adoption of transmission-reducing interventions reduces the prevalence of infected patients. On the other hand, however, the emergence of new cases is extended in time, an effect that produces the desirable trend of “flattening the curve”, ensuring the availability of ICU care for severely ill patients, at the risk of prolonging the scattered emergence of infected patients, eventually giving rise to new waves.

The positive effect of an early lockdown for elderly patients as a single intervention on the evolution of epidemics is clearly visible in our modelling. The role of the elderly in epidemiological dynamics is recognized as the most important factor behind the major COVID-19 outbreak (*Melin et al, 2020*). A number of countries, such as Sweden, adopted a *de-facto* herd immunity approach, without imposing severe limitations to viral propagation in the community, with controversial results (*Bjorklund and Ewing, 2020; Claeson and Hanson, 2020*). The goal of the Swedish Public Health Agency was to assure the intensive care for elderly patients with the highest mortality risk (*Chew et al., 2021*). This approach could probably have been accompanied by a strict lockdown for the elderly. Considering the European incidence of COVID-19 deaths in May 2021, Sweden had a lower per capita incidence of COVID-19 mortality than France, Spain, United Kingdom, Italy, Belgium, or Hungary (*Steward, 2021*), and elderly lockdown is probably one of the most effective interventions to be considered (*Soltesz et al., 2020*).

We should accept that the multifactorial and variable landscape of viral epidemics, where the infection process itself modifies the transmission parameters, is also influenced by the geography, demography, and lifestyles of the population, which makes it difficult to establish general fixed parameters, to a certain extent leading to a “parametric scepticism”. Therefore, the use of flexible models, such as those applied in this study, can mimic differing conditions (that can be adjusted to the locally observed conditions). Integrating parametric ranges and stochastic dynamics is simply a necessity in predicting the policy of corrective interventions in a particular local landscape.

## Data Availability

The data obtained are freely public; in that concernng the accessibility of the model interface, please examine the manuscript.

## Acknowledgements

MC and FB were sponsored by the Projects COV20_00067 of the Program SARS-COV-2 and COVID-19 infection of the Instituto de Salud Carlos III, Ministerio de Ciencia e Innovación of Spain, CB06/02/0053 of the Centro de Investigación Biomédica en Red de Epidemiología y Salud Pública (CIBERESP), and the Regional Government of Madrid (InGeMICS-B2017/BMD-3691). For JCG, this study was partially founded by the Autonomous Community of Madrid, Spain (COVID-19 Grant, 2020) and the Ramón y Cajal Institute for Health Research (IRYCIS), Madrid, Spain. For AM, this study was supported by grants from the Spanish Ministry of Science and Innovation (PID2019-105969GB-I00), the government of Valencia (project Prometeo/2018/A/133) and co-financed by the European Regional Development Fund (ERDF).

## Notes

### Competing Interest Statement

The authors have declared no competing interest.

### Author Declarations

This is an in-silico work, not involving human or animal individuals.

## References

Adam D. Modelling the pandemic: The simulations driving the world’s response to COVID-19. Nature. 2020; 580:316-318.

Ali M., Kim J, Sun-Yoon S. Commensal bacteria in the human intestine produce compounds that inhibit SARS-CoV-2. World Microbe Forum, 2021.

Baquero F. Transmission as a basic process in microbial biology. Lwoff Award Prize Lecture. FEMS Microbiol Rev. 2017. 41:816–827.

Baquero, F. Causality in biological transmission: forces and energies. Microbiol Spectr. 2018, 6(5).

Baquero F, Campos M, Llorens C, et al. 2018. A model of antibiotic resistance evolution dynamics through P systems with active membranes and communication rules. In: Enjoying natural computing. Graciani C, Agustín Riscos-Núñez A, Păun Gh, Rozenberg G, Salomaa A (eds). p 33–44. Springer, Cham. 2018.

Bhatt S. Estimating the effects of non-pharmaceutical interventions on COVID-19 in Europe. Nature. 2020. 584:257–261.

Bjorklund K, Ewing A. The Swedish COVID-19 response is a disaster. It shouldn’t be a model for the rest of the world. Time, Oct 14, 2020. https://time.com/5899432/Sweden-coronovirus-disaster/

Bordia L, Piralla A, Lalle E et al. Rapid and sensitive detection of SARS-CoV-2 RNA using the Simplexa™ COVID-19 direct assay. J Clin Virol. 2020. 128:104416.

Bullard J, Dust K, Funk D, et al. Predicting infectious SARS-CoV-2 from diagnostic samples. Clin Infect Dis. 2020. ciaa638.

Calvo C, López-Hortelano MG, de-Carlos-Vicente JC, et al. Recomendaciones sobre el manejo clínico de la infección por el «nuevo coronavirus» SARS-CoV2. Grupo de trabajo de la Asociación Española de Pediatría (AEP). Anales Pediatr. 2020. 92:241.e1.

Campos M, Llorens C, Sempere JM, et al. A membrane computing simulator of transhierarchical antibiotic resistance evolution dynamics in nested ecological compartments (ARES). Biol Direct. 2015. 10(1), 41.

Campos M, Capilla R, Naya F, et al. Simulating multilevel dynamics of antimicrobial resistance in a membrane computing model. mBio 2019; 10:e02460–18;

Campos M, San Millán A, Sempere JM, et al. Simulating the influence of conjugative plasmids kinetic values on the multilevel dynamics of antimicrobial resistance in a membrane computing model. Antimicrob Agents Chemother. 2020; 64:e00593–20

Casares M, Khan H. A dynamic model of COVID-19: contagion and implications. Carleton Economics Working Papers (CEWP) 2020. 20–02

Chew MS, Blixt PJ, Åhman R, et al. 2021. National outcomes and characteristics of patients admitted to Swedish intensive care units for COVID-19: A registry-based cohort study. Eur J Anaesthesiol. 2021.| 38:335–343.

Claeson M, Hanson S. 2021. COVID-19 and the Swedish enigma. The Lancet 2021. 397: 259–261.

Colson P, Tissot-Dupont H, Morand A, et al. Children account for a small proportion of diagnoses of SARS-CoV-2 infection and do not exhibit greater viral loads than adults. Eur J Clin Microbiol Infect Dis. 2020; 39:1983–1987.

Cox RJ, Brokstad KA. Not just antibodies: B cells and T cells mediate immunity to COVID-19. Nat Rev Immunol. 2020 Oct;20(10):581–582.

Dogra, P., Ruiz-Ramírez, J., Sinha, K., Butner, J. D., Peláez, M. J., Rawat, M., … & Cristini, V. 2020. Innate immunity plays a key role in controlling viral load in COVID-19: mechanistic insights from a whole-body infection dynamics model. medRxiv.

Ferguson N, Laydon D, Nedjati Gilani G, et al. Report 9: Impact of non-pharmaceutical interventions (NPIs) to reduce COVID19 mortality and healthcare demand. 2020

Flaxman S, Mishra S, Gandy A, Unwin HJT, Mellan TA, Coupland H, Whittaker C, Zhu H, Berah T, Eaton JW: Estimating the effects of non-pharmaceutical interventions on COVID-19 in Europe. Nature 2020:1–8

Fonfria, E. S., Vigo, M. I., García-García, D., Herrador, Z., Navarro, M., Bordehore, C. 2020. Essential epidemiological parameters of COVID-19 for clinical and mathematical modeling purposes: a rapid review and meta-analysis. medRxiv.

Fretheim A. The role of children in the transmission of SARS-CoV-2-19 – a rapid review. 2020. Oslo: Folkehelseinstituttet/ Norwegian Institute of Public Health, 2020

Gibney, E. “Whose coronavirus strategy worked best? Scientists hunt most effective policies.” Nature, vol. 581, no. 7806, 2020

Gil-Gil T, Ochoa-Sánchez LE, Baquero F, et al. Antibiotic resistance: time of synthesis in a post-genomic age. Comput Struct Biotechnol J. 2021; 19:3110–3124.

Guan WJ, Ni ZY, Hu Y, et al. Clinical characteristics of coronavirus disease 2019 in China. New Engl J Med. 2020, 220382(18),1708–1720.

Huang AT, Garcia-Carreras B, Hitchings MDT, et al. A systematic review of antibody mediated immunity to coronaviruses: kinetics, correlates of protection, and association with severity. Nat Commun. 2020 Sep 17;11(1):4704)

IHME COVID-19 Forecasting Team. Modeling COVID-19 scenarios for the United States. Nat Med. 2021. 27:94–105.

Kampen J, van de Vijver D, Fraaij P, et al. Shedding of infectious virus in hospitalized patients with coronavirus disease-2019 (COVID-19): duration and key determinants. MedRxiv 2020

Kim YI, Kim SM, Park SJ, et al. Critical role of neutralizing antibody for SARS-CoV-2 reinfection and transmission. Emerg Microbes Infect. 2021. 10.152–160

Kloc M, Ghobrial RM, Kuchar E, et al. Development of child immunity in the context of COVID-19 pandemic. Clin Immunol. 2020. 217:8510

La Scola B, Le Bideau M, Andreani J, et al. Viral RNA load as determined by cell culture as a management tool for discharge of SARS-CoV-2 patients from infectious disease wards. Eur J Clin Microbiol Infect Dis. 2020; 39:1059–1061

Lee B and Raszka WV. COVID-19 Transmission and children: the child is not to blame. Pediatrics. 2020. 146(2):e2020004879;

Lee S, Kim T, Lee E, et al. Clinical course and molecular viral shedding among symptomatic and symptomatic patients with SARS-CoV-2 infection in a community treatment center in the Republic of Korea. JAMA Intern Med. 2020; 180:1447–1452.

Melin M, Ahlbäck Öberg S, Enander A, et al. The Corona Commission. Elderly care during the pandemic. Ministry of Health and Social Affairs, summary of SOU 2020:80. https://www.government.se/4af26a/contentassets/2b394e1186714875bf29991b4552b374/summary-of-sou-2020_80-elderly-care-during-the-pandemic.pdf).

O’Driscoll M, Ribeiro Dos Santos G, Wang L, et al. Age-specific mortality and immunity patterns of SARS-CoV-2. Nature. 2021. 590:140–145.

Oehmke JF, Oehmke TB, Singh LN et al. Dynamic panel estimate-based health surveillance of SARS-CoV-2 infection rates to inform public health policy: model development and validation. J Med Internet Res. 2020; 22(9):e20924.

Păun G, Rozenberg G. Salomaa A (eds). The Oxford Handbook of Membrane Computing. Oxford University Press, Oxford, 2010..

Pérez-Jimenez MJ, Romero-Jimenez A, Sancho-Caparrini F. Complexity classes in models of cellular computing with membranes. Natural Comput. 2003. 2:265–285.

Pierre J. Nudges against pandemics: Sweden’s COVID-19 containment strategy in perspective, Policy and Society. 2020. 39:478–493

Posfay-Barbe K, Wagner N, Gauthey M, et al. COVID-19 in children and the dynamics of infection in families. Pediatrics. 2020. 146(2).

Poland GA. 2020. SARS-CoV-2: a time for clear and immediate action. Lancet Infect Dis., 2020. 20:531–532.

Pujadas E, Chaudhry F, McBride R, et al. SARS-CoV-2 viral load predicts COVID-19 mortality. Lancet Respir Med. 2021; 8:e70.

Singanayagam A, Patel M, Charlett A, et al. Duration of infectiousness and correlation with RT-PCR cycle threshold values in cases of COVID-19, England, January to May 2020. Euro Surveill. 2020. 25:2001483.

Soltesz K, Gustafsson F, Timpka T, et al. The effect of interventions on COVID-19. Nature. 2020, 588: E26–E28.

Sridhar, D, Majumder, D. Modelling the pandemic. BMJ. 2020. 369:m1567

Stewart C. Incidence of coronavirus (COVID-19) deaths in the EEA and the UK 2021, by country (www.statista.com/Statistics/1111779/coronavirus-death-rate-europe-by-country. 2021.

Stokes J, Turner AJ, Anselmi L, et al. The relative effects of non-pharmaceutical interventions on early COVID-19 mortality: natural experiment in 130 countries. medRxiv. 2020.

To KKW, Tsang OTY, Leung WS, et al. Temporal profiles of viral load in posterior oropharyngeal saliva samples and serum antibody responses during infection by SARS-CoV-2: an observational cohort study. Lancet Infect Dis. 2020. 20:565–574.

Van Damme W, Dahake R, van de Pas R et al. 2021. COVID-19: Does the infectious inoculum dose-response relationship contribute to understanding heterogeneity in disease severity and transmission dynamics?. Med Hypoth.2021, 146:110431.

